# Low birthweight neonates and those with long hospital stays are most at risk of antimicrobial-resistant *Klebsiella pneumoniae* infection in Malawi: implications for antibiotic prescribing

**DOI:** 10.64898/2026.06.24.26356242

**Authors:** Tadala Mzengo, Oliver Pearse, Allan Zuza, Mabvuto Chimenya, Jen Cornick, Samantha Lissauer, Chris Jewell, Kondwani Kawaza, Nicholas Feasey

**Affiliations:** Malawi Liverpool Wellcome Research Programme, Kamuzu University of Health Sciences, Blantyre, Malawi; Department of Clinical Sciences, Liverpool School of Tropical Medicine, Liverpool, UK; The School of Medicine, University of St. Andrews, St. Andrews, UK; University of Liverpool, Institute of Infection, Veterinary and Ecological Sciences; Alder Hey Hospital, Liverpool, UK; School of Mathematical Sciences, Lancaster University, Lancaster, UK; Kamuzu University of Health Sciences, Blantyre, Malawi

**Keywords:** sepsis, healthcare-associated infection, neonatal unit

## Abstract

**Research in Context:** *Evidence before this study:* We searched PubMed for studies published between Jan 1, 2015, and Dec 31, 2025, using combinations of the terms “*Klebsiella pneumoniae*”, “neonate”, “young infant”, “bloodstream infection”, “sepsis”, “risk factors”, and “sub-Saharan Africa”. Only seven studies were identified. These studies were identified in South Africa, Tanzania, and Ethiopia. They have identified Gram-negative bacteria, including *Klebsiella pneumoniae*, as important causes of neonatal bloodstream infections and mortality. Previous work has described the epidemiology, antimicrobial resistance profiles, and outcomes of neonatal bloodstream infections, highlighting the increasing burden of multidrug-resistant and extended-spectrum β-lactamase-producing organisms. Studies have also identified risk factors for neonatal sepsis, including prematurity, low birthweight, healthcare exposure, and invasive procedures. However, no studies in Sub-Saharan Africa have specifically examined risk factors for Klebsiella pneumoniae infection among infants younger than 3 months of age. Furthermore, most available studies have focused on clinical outcomes, antimicrobial resistance, or transmission pathways rather than determinants of Klebsiella pneumoniae infection. Data from sub-Saharan Africa remain limited, particularly for young infants in high-burden settings. To our knowledge, no validated risk prediction tools are available to identify infants at increased risk of Klebsiella pneumoniae infection, and there is limited evidence linking risk stratification approaches to targeted treatment or management strategies. No such data have been reported from Malawi.

*Added value of this study:* This study investigated risk factors associated with *Klebsiella pneumoniae* infection among infants younger than 3 months of age in Malawi. We identified factors independently associated with *Klebsiella pneumoniae* infection in early infancy, and we quantified the risk of neonates getting these *Klebsiella pneumoniae* infections, and this was done in a setting where rapid microbiological diagnosis is often unavailable. We also linked risk prediction to potential clinical management by evaluating antibiotic susceptibility patterns and identifying antibiotic regimens that may be appropriate for infants classified as being at high risk of *Klebsiella pneumoniae* infection. Our findings provide evidence from a high-burden African setting where data on pathogen-specific risk prediction and treatment guidance remain scarce.

*Implications of all the available evidence:* Together with existing evidence, our findings provide further evidence that neonatal sepsis in African settings is commonly healthcare associated and antimicrobial resistant. Antimicrobial stewardship is not simply about restriction to unnecessary therapy, but also enabling access where it has the potential to be life-saving. Neonates do not have the physiological reserve to wait for antimicrobial therapy to be tailored to a blood culture result. Early identification of infants with clinical suspicion of sepsis that are at increased risk of *Klebsiella pneumoniae* infection could support more targeted clinical management and reduce mortality due to *Klebsiella pneumoniae* and improve antimicrobial stewardship in high-burden settings.

**Background:** *Klebsiella pneumoniae* (*Kpn*) is a major cause of neonatal sepsis in Africa. 3^rd^-generation cephalosporin and gentamicin resistant *Kpn* is the norm in many sites, rendering WHO-recommended first- and second-line antimicrobials ineffective. An understanding of which neonates and infants are most at risk of sepsis caused by *Kpn* would support the case for improved access to WHO watch and reserve antimicrobials (i.e. carbapenems) for patients most likely to benefit from them.

**Methods:** A prospective case-control study was conducted at Queen Elizabeth Central Hospital, Malawi. Cases were infants <3 months of age with blood or CSF culture-confirmed Kpn infection. Controls were healthy infants from the same wards and were matched 2:1. Univariate and multivariate logistic regression were performed on mean-centred data to determine risk factors for infection with Kpn.

**Results:** We analysed data from 38 cases and 76 controls between [insert dates]. Mortality at 3 months of age was 21/38 (29%) for cases, with 14/38 (37%) identified postmortem and 6/76 (7.9%) for controls (OR 14.0 (95% CI 4.59, 49.2, p>0.001). Cases were more likely to be born out of QECH than controls (42% vs. 24%, p = 0.043), and cases had lower birthweights (median 2200g vs. 2850g, p = 0.005). Multivariate logistic regression analysis revealed that increasing birthweight was protective against *Kpn* infection (OR: 0.858 [95% CI: 0.745–0.987] per 100g increase), while longer hospital stay was associated with increased odds of infection (OR: 1.148 [95% CI: 1.012–1.1.303] per additional day). Most infecting isolates (34/38 [89%]) were resistant to first- and second-line antimicrobial agents, but all were sensitive to meropenem and 33/36 [92%] to amikacin.

**Conclusion:** Low birthweight infants with prolonged hospital stay were at greatest risk of *Kpn* infections that were typically resistant to WHO first- and second-line antimicrobial therapy. These infants should be prioritised for antibiotics that have the potential to be life-saving. The overlapping and evolving nature of these risk factors makes it difficult to design a simple tool to support empiric initiation of meropenem. Neonates critically ill with *Kpn* sepsis cannot, however, afford to wait for blood culture-confirmation before receiving effective treatment. This highlights the need for empiric decision-making frameworks that allow rapid initiation of effective therapy in high-risk neonates.

## Introduction

Sustainable Development Goal 3.2 is to eliminate preventable deaths among neonates, with a focus on reducing neonatal mortality rates to 12 or less per 1,000 live births by 2030 (1). *Klebsiella pneumoniae* (*Kpn*) is a leading cause of neonatal infection globally (2,3)leading to substantial morbidity and mortality. Increasing antimicrobial resistance (AMR) in *Kpn* means that isolates in many locations are typically resistant to first- and second-line antibiotics. This problem is further exacerbated in Malawi and other Sub-Saharan African countries where access to antimicrobials is limited such that these infections can be impossible to treat (4).

First-line treatments for neonatal sepsis and meningitis in Malawi and many other countries follow World Health Organization (WHO) guidelines recommending penicillin and gentamicin in combination (5). Second-line therapy often includes a third-generation cephalosporin (3GC) such as ceftriaxone, however where blood culture is available, laboratory data suggest ceftriaxone may add little to this regimen (6,7). In ceftriaxone and gentamicin-resistant infections (*Kpn* is intrinsically resistant to aminopenicillins), which are the norm in many locations, carbapenems such as meropenem or aminoglycosides such as amikacin are required to treat neonatal infection, though these are expensive and may not be routinely available (6,8).

It typically takes 3-5 days from the point at which a blood culture is taken to provide a final result including isolate identification and antimicrobial susceptibility testing and in many settings these tests are not available (9,10). In neonatal sepsis, however, best practice dictates that antibiotics should be administered as early as possible, with international guidance recommending treatment within one hour of the decision to treat to optimise outcomes (11). Delays in initiating effective antimicrobial therapy are associated with increased mortality, particularly in severe infections (12). Thus, by the time a blood culture result has been returned, patients with *Kpn* infection may be either too unwell to benefit from an antimicrobial change or already dead (12).. In absolute terms, the proportion of neonates presenting with suspected infection who have 3GC resistant *Kpn* is also low (13), therefore despite the high likelihood of treatment failure with first- or second-line antibiotics for *Kpn* infection it is not feasible to give all neonates presenting with suspected infection carbapenems or amikacin due to their cost, poor availability, and the risk of driving increases in AMR In areas with a high prevalence of 3GC resistant *Kpn*, simple decision-support tools to determine which neonates and infants are at greatest risk of *Kpn* infection are needed to facilitate access to extended-spectrum beta-lactamase producing (ESBL) stable antimicrobials for empiric treatment of at-risk patients early in the course of their infection. This requires determination of risk factors that predispose neonates to infection with *Kpn.* By identifying the most important risk factors, healthcare providers can tailor empiric antibiotic therapies to at-risk individuals, ultimately mitigating the morbidity and mortality associated with AMR infections. In this study we aimed to describe risk factors for invasive *Kpn* infection at QECH, Malawi, as a first step in establishing decision-support tools in antimicrobial prescribing for neonates and infants.

## Methodology

### Study location

This study was conducted on two wards at Queen Elizabeth Central Hospital (QECH): Chatinkha nursery and paediatric nursery. QECH is the largest government-run referral and teaching hospital in southern Malawi, providing healthcare free at the point of delivery. Chatinkha nursery is the neonatal unit at QECH. There are approximately 14,000 neonates born at QECH each year and 5,000 admissions to the Chatinkha nursery. Approximately 60% of neonates are born inside QECH and the rest are born at other facilities or at home (14). There are typically between 30 and 70 neonates on the ward at any one time. Any neonate born at QECH with complications during or after birth is admitted for observation and/or management, as well as infants with complex medical or surgical needs from other hospitals; almost none of the neonates admitted to Chatinkha nursery have ever been out of hospital. The paediatric nursery admits infants up to 6 months of age that have been discharged from hospital and been admitted via Accident and Emergency (A and E). Infants in both wards can receive oxygen therapy, continuous positive airway pressure, intravenous medication, blood transfusion, radiological investigations, blood tests and nasogastric feeding, neither has intensive care facilities.

### Recruitment

This case-control study was nested within a prospective observational cohort at QECH, where infants ≤3 Months-old admitted to the Chatinkha or paediatric nursery at QECH were recruited (Figure 1A) (15). We also recruited mothers in labour who were expected to deliver at QECH and followed their neonates to 3 months post-delivery. Cases were infants with culture-confirmed infection with *Kpn* in a normally sterile site (blood or Cerebral Spinal Fluid (CSF)). Some cases were recruited posthumously, with data collected retrospectively from their patient records (ethics was granted to collect data from these neonates without informed consent). Controls were infants with no clinical features of infection, less than or three months of age, from the same wards as the cases. The criteria for infection were: documented fever ≥ 38℃, clinical diagnosis of sepsis or meningitis as per the clinical team, blood culture or lumbar puncture performed. Data were collected using Open Data Kit (ODK) on study tablets, anonymised and only accessible to trained study team members and stored on a password-protected server at the Malawi Liverpool Wellcome Research Programme (MLW) (16). For all mother-infant pairs, stool samples or rectal swabs were collected from both the mother and infant, using sterile non-touch technique. Stool samples were stored in universal containers. Rectal swabs were stored in Amies media.

**Figure 1:**
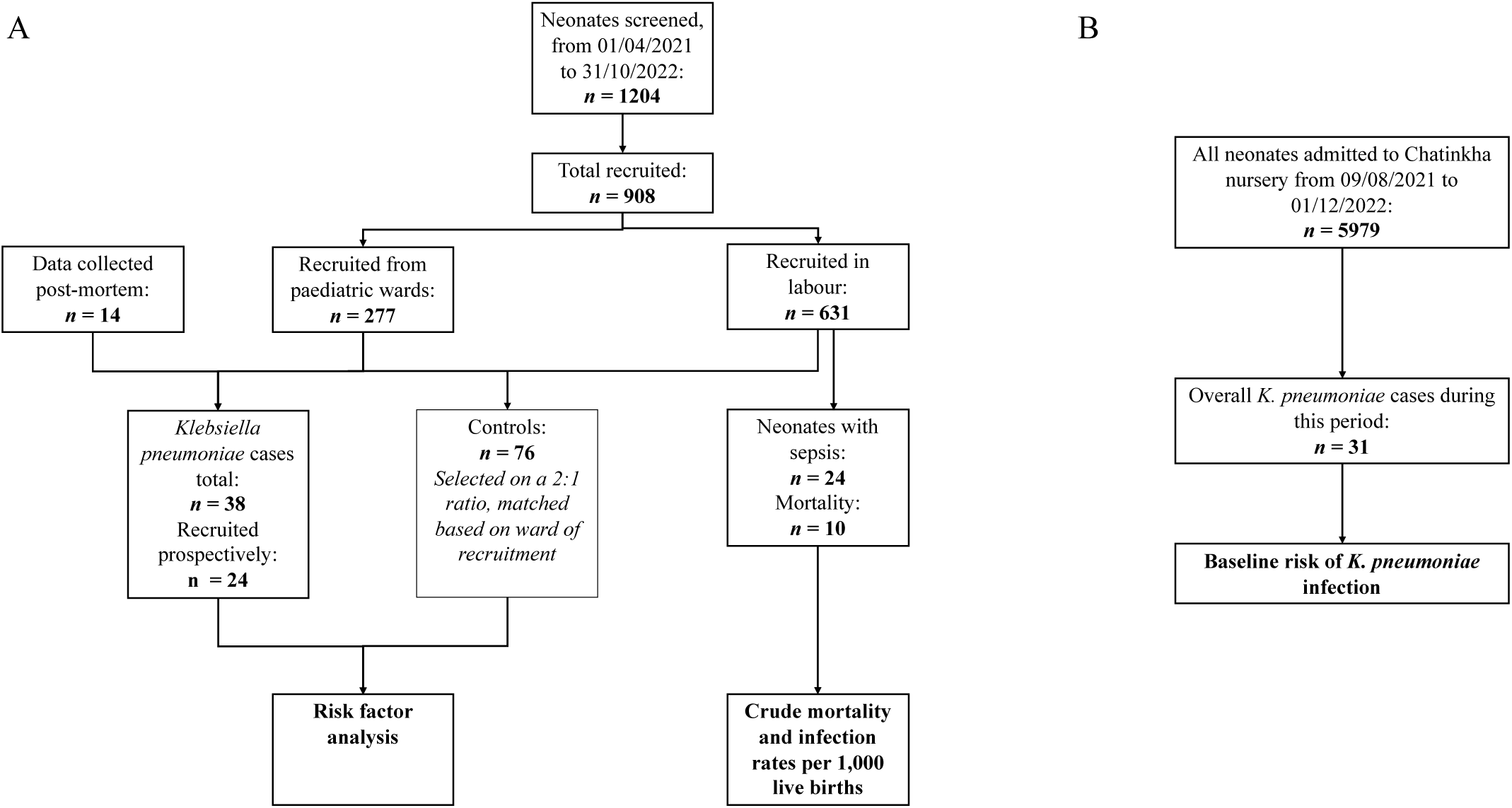
A flow chart detailing patient recruitment and the sources of data for the case-control risk factor analysis and calculation of the crude mortality and infection rates per 1,000 live births. B) A flow chart detailing the sources of data used to calculate the baseline risk of Kpn infection.

### Laboratory processing

Routine, quality-assured diagnostic blood culture services have been provided to children with features of infection by the ISO15189 accredited MLW diagnostic microbiology laboratory since 1998. Briefly, 1-2mL of blood was taken from infants with risk factors for sepsis (i.e. maternal fever during labour, prolonged rupture of membranes), or clinical suspicion of sepsis (fever >38℃, tachypnoea, tachycardia, reduced activity, seizures). For neonates with clinical suspicion of sepsis or other clinical suspicion of meningitis (raised fontanelle, reduced level of consciousness, abnormal neurology), a lumbar puncture was also performed and CSF sent for culture. Blood was collected using aseptic methods and inoculated into a single aerobic bottle (BacT/Alert, bioMérieux, Marcy-L’Etoile, France), then incubated using the automated BacT/Alert system. Samples that flagged positive were Gram-stained. Gram-negative bacilli were identified by Analytical Profile Index (bioMérieux). Antimicrobial susceptibility testing was determined by the disc diffusion method (Oxoid, United Kingdom) according to European Committee on Antimicrobial Susceptibility Testing (EUCAST) guidelines. *Kpn* isolates were stored at −80°C on microbank beads.

Stool samples and rectal swabs were streaked onto Uriselect agar and incubated for 18 - 24 hours at 37℃ (17). Blue colonies on Uriselect agar were identified as potential *Kpn* and subcultured before being further identified using melt-curve PCR analysis as previously published (18). *Kpn* isolates were stored at −80°C in skim milk, tryptone glucose and glycerin (STGG) (18).

### Statistical analysis

R programming software was used for statistical analysis (19) and data visualisation. The Kolmogorov–Smirnov test and visual inspection was used to evaluate normality of the data. Non-normally distributed variables were reported as medians with interquartile ranges (IQRs) and compared using the Wilcoxon rank-sum test for pairs or median test for multiple groups. Differences in proportions among categorical data were assessed using Fisher’s exact test for pairwise comparisons and the Pearson’s Chi Squared for multiple groups.

To determine the overall 3-month crude mortality rate and infection rate of neonates born at our site, data from the cohort of mothers that were recruited in labour was analysed. Within this cohort the outcomes of death and infection per 1,000 live births were calculated as described in the supplementary methods (Figure 1A; Section S2).

Variables were centred to the mean prior to model fitting. Missing data was imputed, with 20 imputations performed and the results combined, using the MICE v3.17.0 package in R(19,20). For both univariate and multivariate models, risk factors affecting *Kpn* infection were analysed using logistic regression. This was performed utilising the glm function in base R, specifying a binomial family with a logit link function. Risk factors included in the models were neonatal colonization with *Kpn*, mode of delivery, maternal colonization with *Kpn*, time in hospital, Human Immunodeficiency Virus (HIV) status, gestational age, gender, birthweight and birthplace. Time in hospital was defined as described in the Supplementary Methods (Section S1).

Receiver operating characteristic (ROC) curves were generated to evaluate the discriminative performance of logistic regression models predicting *Kpn* infection. Area under the curve (AUC) values were calculated for the full model, including all covariates, as well as for a reduced model in which non-significant covariates were set to their mean values. ROC curves are presented in Supplementary Figure S1.

We calculated the probability of *Kpn* infection for neonates admitted to the Chatinkha nursery utilising the number of culture-confirmed *Kpn* infections on the nursery over the period from August 2021 to April 2023 and the number of admissions to Chatinkha nursery from the same period (Figure 1B). This was used as the baseline probability of *Kpn* infection, to which the OR was applied to determine the increased probability of low-birthweight infants admitted to the ward for three weeks. This is further explained in the supplementary methods (Section S4).

### Ethical requirements

Ethical approval for the study was granted by the local ethics board COMREC (study number P.10/18/2499) and LSTM REC (study number 19-018). LSTM acted as the study sponsor.

## Results

We screened 1204 participants for inclusion, and 908 were recruited into the study. From these a total of 38 culture-confirmed cases were described, 24 prospectively recruited and 14 post-mortem identified cases, with 76 controls (Figure 1). Mortality at 3 months of age was 21/38 (29%) for cases, with 14/38 (37%) recruited postmortem and 6/76 (7.9%) for controls (OR 14.0 (95% CI 4.59, 49.2, p>0.001). In the live birth cohort, there were 10 deaths and 24 cases of clinically defined sepsis by three months from 631 live births, giving a crude mortality rate of 15.8 per 1,000 live births (95% CI: 7.6 – 28.1) and a crude infection rate of 38.0 per 1,000 live births (95% CI: 24.4 – 56.6). 93/631 (14.7%) of neonates were admitted to one of the two neonatal and infant wards, of which 89/93 (96%) were admitted directly or soon after labour without being discharged and 4/93 (4%) were readmitted from home.

Median birthweight was lower in cases (2,200g [1,650.00, 2,900.00]; Table 1) than in controls (2,850g [2,200.00, 3,200.00] *p*=0.005). Length of hospital stay was longer for cases (median stay 4 days [1.00, 8.00]) than for controls (median stay of 1 day [0.00,3.00]), *p*=0.007). Cases were more likely than controls to be born outside of QECH (42% vs. 24%, p = 0.043) (Table 1).

**Table 1:** Clinical characteristics of cases and controls.

| Characteristic | Controls<br>N = 76 | Cases<br>N = 38 | *p*-value |
| --- | --- | --- | --- |
| Gestational age in weeks, median [Q1 - Q3] | 38.00 (35.00, 39.00) | 36.50 (31.50, 38.00) | 0.071<sup>1</sup> |
| Birthweight in grams, median [Q1 - Q3] | 2,850.00 (2,200.00, 3,200.00) | 2,200.00 (1,650.00, 2,900.00) | **0.005**<sup>1</sup> |
| HIV status, Positive n(%) | 10 (14%) | 4 (11%) | 0.80<sup>2</sup> |
| Mode of delivery, SVD, n(%) | 60 (79%) | 28 (78%) | 0.50<sup>3</sup> |
| Maternal Colonization at recruitment, n(%) | 7 (9.2%) | 4 (11%) | >0.90<sup>3</sup> |
| Infant colonization at recruitment, n(%) | 7 (9.2%) | 9 (24%) | **0.031**<sup>3</sup> |
| Age at recruitment in days, Median [Q1 - Q3] | 1.50 (0.00, 4.00) | 8.00 (4.50, 14.50) | <**0.001**<sup>1</sup> |
| Birthplace, n(%) |  |  | **0.043**<sup>2</sup> |
| Outside Queens | 18 (24%) | 16 (42%) |  |
| Inside Queens | 58 (76%) | 22 (58%) |  |
| Gender, Male, n(%) | 40(54%) | 17 (52%) | >0.80<sup>2</sup> |
| Length of Hospital Stay, Median [Q1 - Q3] | 1.00 (0.00,3.00) | 4.00 (1.00, 8.00) | **0.007**<sup>1</sup> |
| Died | 6 (7.9%) | 21 (55%) | <**0.001**<sup>2</sup> |

<sup>1</sup> Wilcoxon rank sum test
<sup>2</sup> Pearson's Chi-squared test
<sup>3</sup> Fisher's exact test

### Antimicrobial susceptibility of cultured isolates

All isolates were resistant to ampicillin (*Kpn* has constitutive ampicillin resistance). There was also high prevalence of gentamicin resistance (33/38 [87%]) and ceftriaxone resistance 34/38 [89%]). In contrast, there was no meropenem resistance and infrequent amikacin resistance (3/36 [8%]). 33/38 (87%) of isolates were resistant to first-line (ampicillin and gentamicin) therapy, and 32/38 (84%) of isolates were resistant to both first-line and second-line therapy (ceftriaxone). Isolates were commonly resistant to co-trimoxazole (35/38 [92%]) and piperacillin/tazobactam (26/38 [72%]) but ciprofloxacin (20/38 [53%]) and chloramphenicol (16/38 [42%]) resistance was less common (Figure 2).

**Figure 2:**
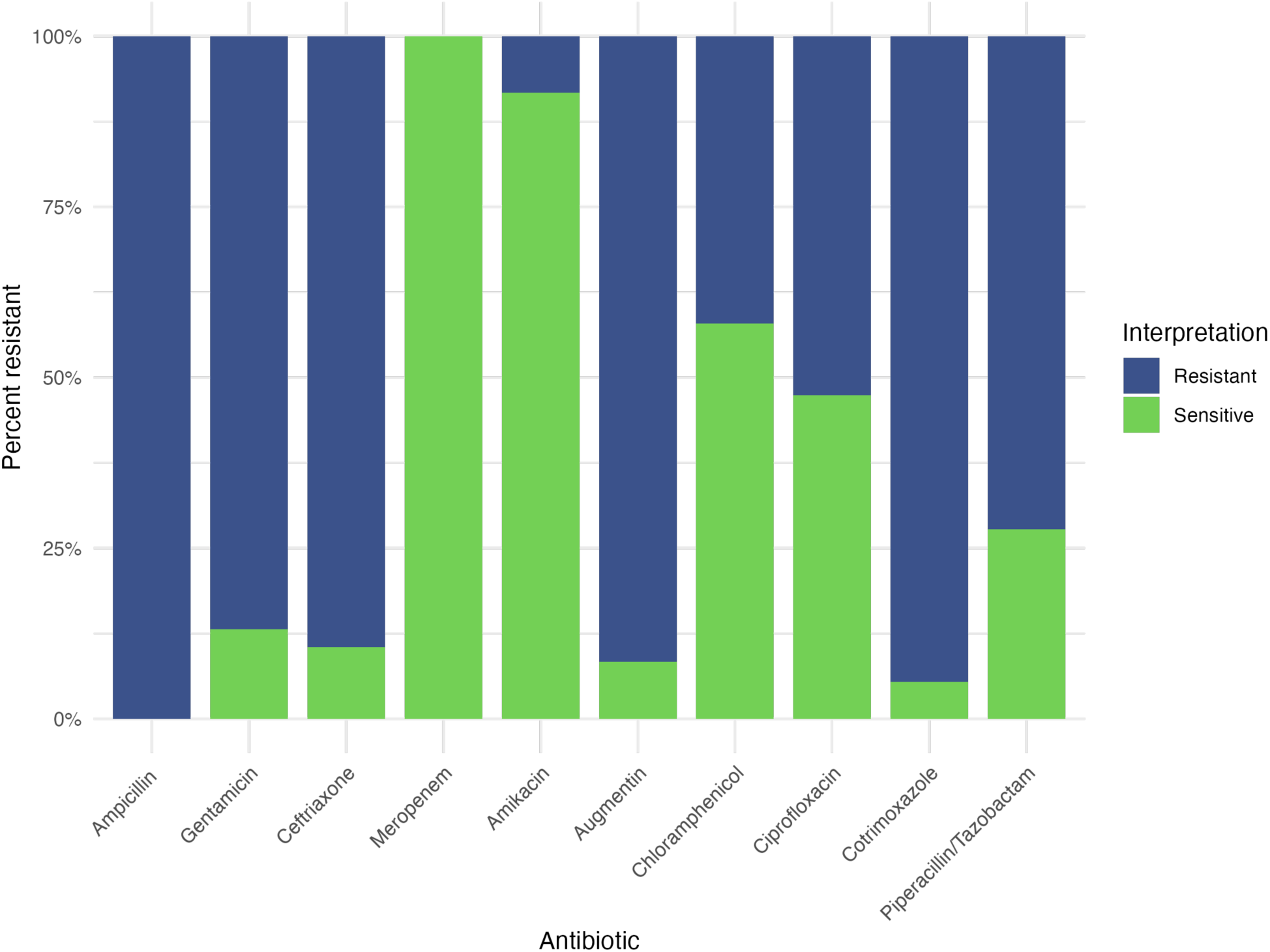
Antibiotic resistance patterns from neonatal sepsis Klebsiella isolates.

### Risk factors for *Kpn* infection

In both univariate and multivariate analyses, two factors were significantly associated with *Kpn* infection: birthweight and length of hospital stay (Table 2). Specifically, in the univariate analysis, a 100-gram increase in birthweight was associated with an odds ratio (OR) of 0.921 (95% CI 0.868 – 0.977), and in the multivariate analysis an OR of 0.858 (95% CI: 0.745 – 0.987) per 100-gram increase, meaning that after adjusting for other variables, each 100-gram increment in birthweight decreased odds of infection by approximately 14.2%. Each additional day in hospital increased the odds of *Kpn* infection in the univariate analysis (OR = 1.186; 95% CI: 1.054 –1.335) and the multivariate analysis (OR = 1.012; 95% CI: 1.012–1.303). Additionally, the univariate analysis identified two other risk factors: infant colonization (OR = 3.168; 95% CI: 1.054–9.346), and Birth inside QECH with lower odds of Kpn infection (OR 0.43, 95% CI 0.19–0.98). This association was similar in magnitude after adjustment in the Multivariate analysis, although the confidence interval crossed unity (OR 0.41, 95% CI 0.15–1.11) (Table 2).

**Table 2:** Univariate and multivariate logistic regression analyses identifying risk factors associated with Kpn infection.

|  | Univariate Analysis |  |  |  | Multivariate Analysis |  |  |  |
| --- | --- | --- | --- | --- | --- | --- | --- | --- |
|  | Odds Ratio | 95% CI |  | P-value | Odds ratio | 95% CI |  | P-value |
|  |  | Lower | Upper |  |  | Lower | Upper |  |
| **Birthweight** | **0.921** | **0.868** | **0.977** | **0.007** | **0.858** | **0.745** | **0.987** | **0.037** |
| **Maternal colonization at recruitment** | 1.160 | 0.318 | 4.24 | 0.823 | 0.320 | 0.054 | 1.860 | 0.207 |
| **Gestational age** | 0.923 | 0.823 | 1.035 | 0.1734 | 1.215 | 0.931 | 1.587 | 0.157 |
| **Infant colonization at recruitment** | **3.168** | **1.074** | **9.346** | **<0.039** | 2.854 | 0.770 | 10.566 | 0.119 |
| **Length of Hospital Stay** | **1.186** | **1.054** | **1.335** | **0.005** | **1.148** | **1.012** | **1.303** | **0.033** |
| **Mode of delivery (SVD)** | 1.161 | 0.431 | 3.127 | 0.768 | 1.367 | 0.405 | 4.614 | 0.615 |
| **Gender (male)** | 1.138 | 0.502 | 2.576 | 0.757 | 1.137 | 0.505 | 3.927 | 0.514 |
| **HIV Status (positive)** | 0.631 | 0.192 | 2.077 | 0.450 | 0.599 | 0.154 | 2.323 | 0.460 |
| **Birthplace (inside QECH)** | **0.427** | **0.185** | **0.982** | **0.048** | 0.406 | 0.149 | 1.110 | 0.08 |

### Risk Prediction of Kpn

We used these data to compare odds of *Kpn* infection in an infant born at a healthy birthweight (2600g, 2586g is the mean birthweight in the labour cohort) and discharged one day after birth to a low-birthweight preterm infant (1500g) who has a lengthy hospital stay (21 days). Baseline probability of *Kpn* infection on the Chatinkha nursery was 0.52% (95% CI: 0.35 – 0.74) over the period from August 2021 – April 2023, though this varies significantly depending on whether there is an outbreak occurring.

To determine the extent to which these variables predicted *Kpn* infection we created ROC curves and calculated the AUC. The full model demonstrated good discrimination with an AUC of 0.80 and when only birthweight and length of time in hospital was included (by fixing other covariates at their mean values) the AUC decreased to 0.72, indicating that the model’s predictive performance was primarily but not completely driven by these covariates (Supplementary Figure S1).

Figure 3 shows a heatmap illustrating how the predicted probability of *Klebsiella* infection increases with longer hospital stays and lower birthweights. Neonates with both low birthweights and extended hospital stays display highest infection risks. In contrast, neonates with higher birthweights and shorter hospital stays show a substantially lower predicted risk.

**Figure 3:**
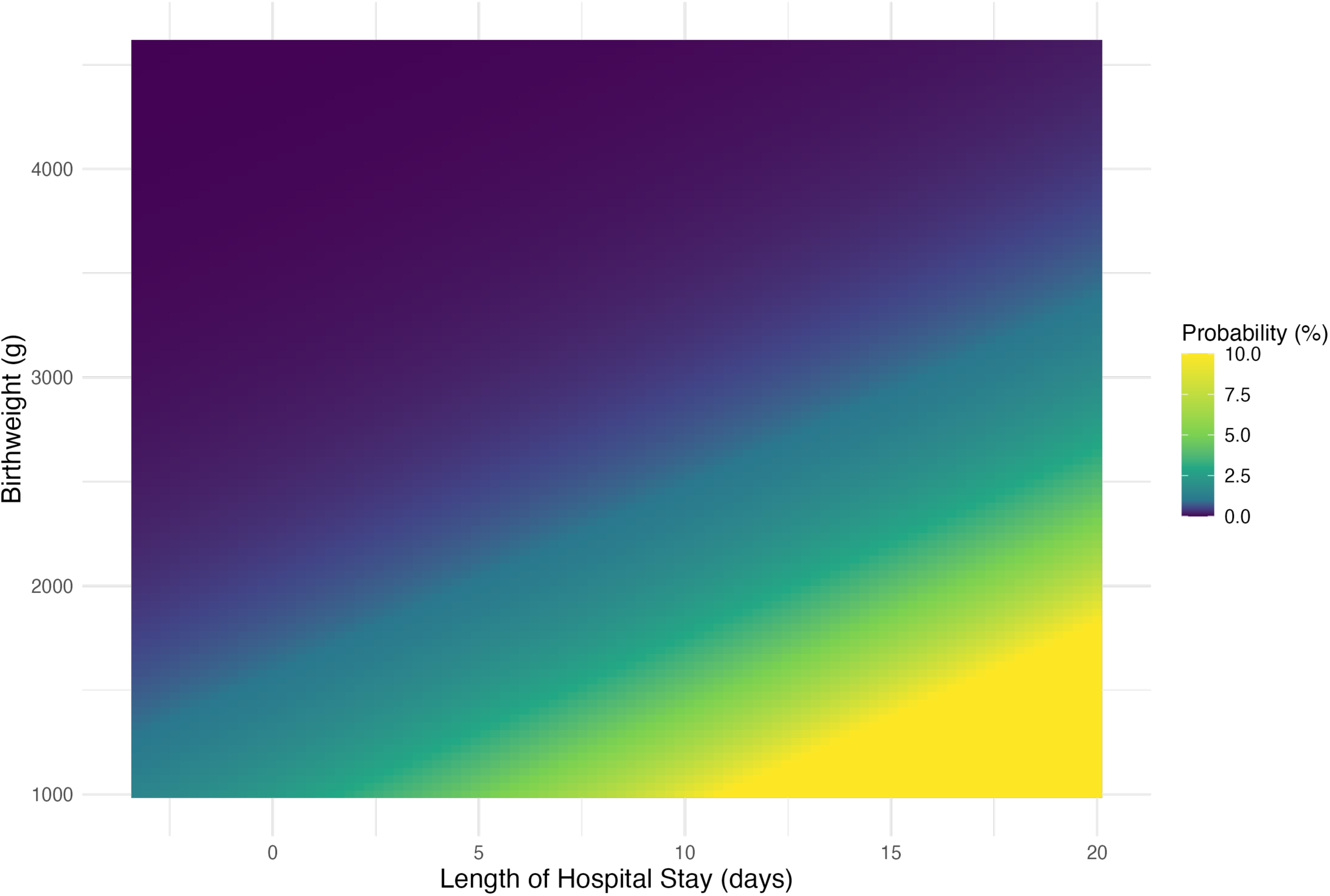
Klebsiella pneumoniae neonatal sepsis risk prediction heatmap.

## Discussion

This study describes clinical characteristics of infants less than three months old infected with *Kpn* in a Malawian hospital, revealing high mortality from sepsis caused by *Kpn*. We reveal that low birthweight and increasing length of hospital stay are key risk factors for *Kpn* infection and that in plausible clinical scenarios, these risk factors multiply, meaning that low-birthweight infants (which often have to be admitted for long periods of time) become highly vulnerable to infection with *Kpn* and should be prioritised for access to empiric watch or reserve antibiotics (in this setting meropenem) if they become unwell.

The results of our study are consistent with studies conducted in other locations (21–23). Length of stay in hospital was a risk factor for infection, which suggests that these are likely healthcare-associated infections (HAIs) rather than transmitted in the birth canal. Prolonged hospitalisation increases risk of exposure to bacteria that thrive in the healthcare environment, thereby elevating the likelihood of HAIs. A retrospective cohort study focusing on premature infants found extended hospitalisation was associated with higher incidence of *Kpn* infections (21). Similarly, research on carbapenem-resistant *Kpn* (CRKP) outbreaks in neonatal intensive care units (NICUs) have highlighted prolonged hospitalisation as a risk factor (22).

Vulnerability of low birthweight infants to *Kpn* infection is likely due to their inherent vulnerability to any infection, the necessity for more invasive interventions (such as IV medications, respiratory support and feeding support), their higher likelihood of receiving antimicrobial therapy and longer hospital stays compared to their normal birthweight counterparts (23). Our findings underscore the importance of recognizing and addressing the unique challenges faced by low birthweight infants, who require heightened vigilance in healthcare settings to mitigate the risk of all invasive infections but particularly *Kpn*. In general prematurity is often a better indicator for clinical vulnerability than birthweight and as a result would potentially be expected to have a greater effect on *Kpn* infection than birthweight. However, in Malawi the estimation of gestational age is not always accurate, and in this context, birthweight may be a more accurate surrogate marker of gestational age than the documented gestational age.

Certain factors which were related to *Kpn* infection in the univariate model were lost in the multivariate model (neonatal colonisation with *Kpn* and being born outside QECH), which might be because both factors are proxies for the true exposures which are length of stay in hospital (neonates are only transferred to QECH if they are significantly unwell and requiring transfer to a referral hospital) and low birthweight. This may also reflect small sample size.

The risk prediction heatmap helps visualise interaction of low birthweight and prolonged hospitalisation in elevating likelihood of *Kpn* infection. It is illustrative of one setting and not designed to guide individual treatment decisions, but the figure helps clarify why certain neonates may warrant early escalation to broader-spectrum (i.e. ESBL-stable) antibiotics. It helps communicate risk patterns in a clinically intuitive way, supporting discussions around empiric antibiotic use in high-risk subgroups. However, this simplifies a complex clinical reality and cannot account for real-time changes in an infant’s condition or external factors like unit-level transmission dynamics.

Prevalence of AMR to first- and second-line antibiotics in the isolates in this study were very high. These findings are consistent with results from other studies. Similarly, the CHAMPS (Child Health and Mortality Prevention Surveillance) study, which conducted post-mortem investigations into causes of child mortality in multiple African and Asian countries, reported that their findings highlighted the inadequacy of current first- and second-line empirical treatments, contributing to high mortality rates in neonates (2,6,24).

We found high mortality amongst *Kpn* cases compared to controls. This high mortality may partly reflect delays in initiation of effective antimicrobial therapy, as most neonates are receiving an ineffective antibiotic due to resistance (11). This highlights the significant burden of *Kpn* infections on patient outcomes. Our findings highlight the infants most at risk from *Kpn* infection and those which would most benefit from tailored antibiotic therapy. In our study, the archetypal infant that became infected with *Kpn* would be a low-birthweight infant with a resultant prolonged hospital stay. These findings should be applicable to other settings which have similar resource constraints. Based on our findings, consideration should be given for empiric administration of antibiotics such as meropenem or amikacin to low-birthweight infants that have been on the ward for some time that become clinically septic. Some high-income countries, such as the UK, already tailor empiric antibiotic regimens based on the timing of suspected neonatal sepsis (early-onset usually defined as ≤48–72 hours vs. late-onset thereafter) (25).

In addition to the effect of these findings on antimicrobial prescribing policy, they also have implications for infection prevention and control (IPC). Healthcare practitioners should be particularly attentive to infection prevention and control measures to prevent nosocomial transmission of bacteria to safeguard the most vulnerable in this generally fragile patient population. Additionally, efforts to minimise the length of hospital stays for neonates, when medically appropriate, may contribute to reducing the incidence of hospital-acquired *Kpn* infections.

This study had several limitations. Firstly, the number of culture-confirmed *Kpn* cases was relatively small, reducing the statistical power and precision in estimating associations. These limitations highlight the need for larger studies to validate and expand on these findings.

### Conclusion

*Kpn* infection in Malawi has high mortality. Low birthweight infants and those with prolonged admissions are particularly at risk and consideration should be given to change antibiotic prescribing policy such that these neonates are offered empiric antibiotic therapy that is likely to be active against locally circulating bacteria as soon as they develop clinical signs of infection. This approach is consistent with best practice in management of neonatal sepsis globally. A more targeted approach that prioritises early escalation to meropenem in neonates with overlapping risk factors, such as low birthweight and prolonged hospitalisation, is warranted, even in the absence of microbiological confirmation.

## Data Availability

All data produced in the present study are available upon reasonable request to the authors.

## Supplementary methods

### S1. Definition of Time in Hospital

Time in hospital was defined as the total number of days an infant had spent in hospital during the relevant admission period prior to study inclusion. For cases, this was calculated up to the date of blood culture collection, confirming *Kpn* infection. For controls, time in hospital was calculated up to the date of recruitment. For infants born at Queen Elizabeth Central Hospital (QECH) or transferred there directly from another healthcare facility, time in hospital represented the continuous duration from birth to the recruitment date. For infants who had been discharged home and subsequently readmitted, time in hospital was defined as the number of days since their most recent return to hospital, including time spent at QECH and any other healthcare facilities during that admission.

### S2. Crude Mortality and Infection Rate Calculations

Crude mortality rates per 1,000 live births, overall infection rates per 1,000 live births, and *Kpn* infection rates per 1,000 hospital admissions were calculated. Event counts were assumed to follow a Poisson distribution, and 95% confidence intervals were derived using the chi-squared distribution.

Rates and corresponding confidence intervals were calculated as follows:

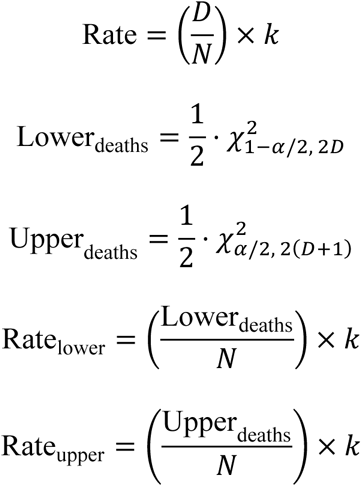

Where *D* = number of events, *N* = number of live births/admissions, *k* = scale factor (in this case 1,000) and α= 0.05.

### S3. Absolute Risk Estimation and Heatmap Construction

To examine the joint effect of birthweight and length of hospital stay on the risk of *Kpn* infection, absolute risk estimates were generated and visualised using a heatmap. A baseline probability of infection was first estimated for a reference neonate, defined as having the mean birthweight and the mean length of hospital stay within the study cohort. This baseline probability was converted to baseline odds using the relationship:

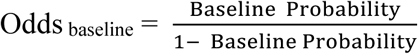

Birthweight and length of hospital stay were then expressed as deviations from the reference values. Birthweight was scaled per 100 g difference from the cohort mean (2586g), and length of stay was defined as days since admission relative to one day:

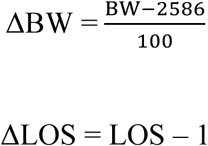

The combined effect of birthweight and length of stay was calculated by exponentiating the corresponding odds ratios from the regression model:

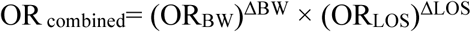

where OR_BW_ represents the odds ratio per 100 g increase in birthweight and OR_LOS_ represents the odds ratio per additional day of hospital stay.

New odds were obtained by multiplying the baseline odds by the combined odds ratio, and these were converted back to probabilities:

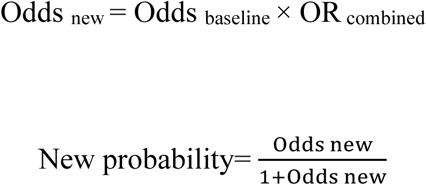

Predicted probabilities were calculated across a grid of birthweights ranging from 1,000 g to 4,000 g and lengths of hospital stay ranging from 0 to 40 days. At each grid point, the baseline probability was adjusted according to the cumulative relative effects of birthweight and length of stay, and the resulting estimates were used to construct the heatmap.

## Supplementary figures and tables

**Supplementary Figure S1:**
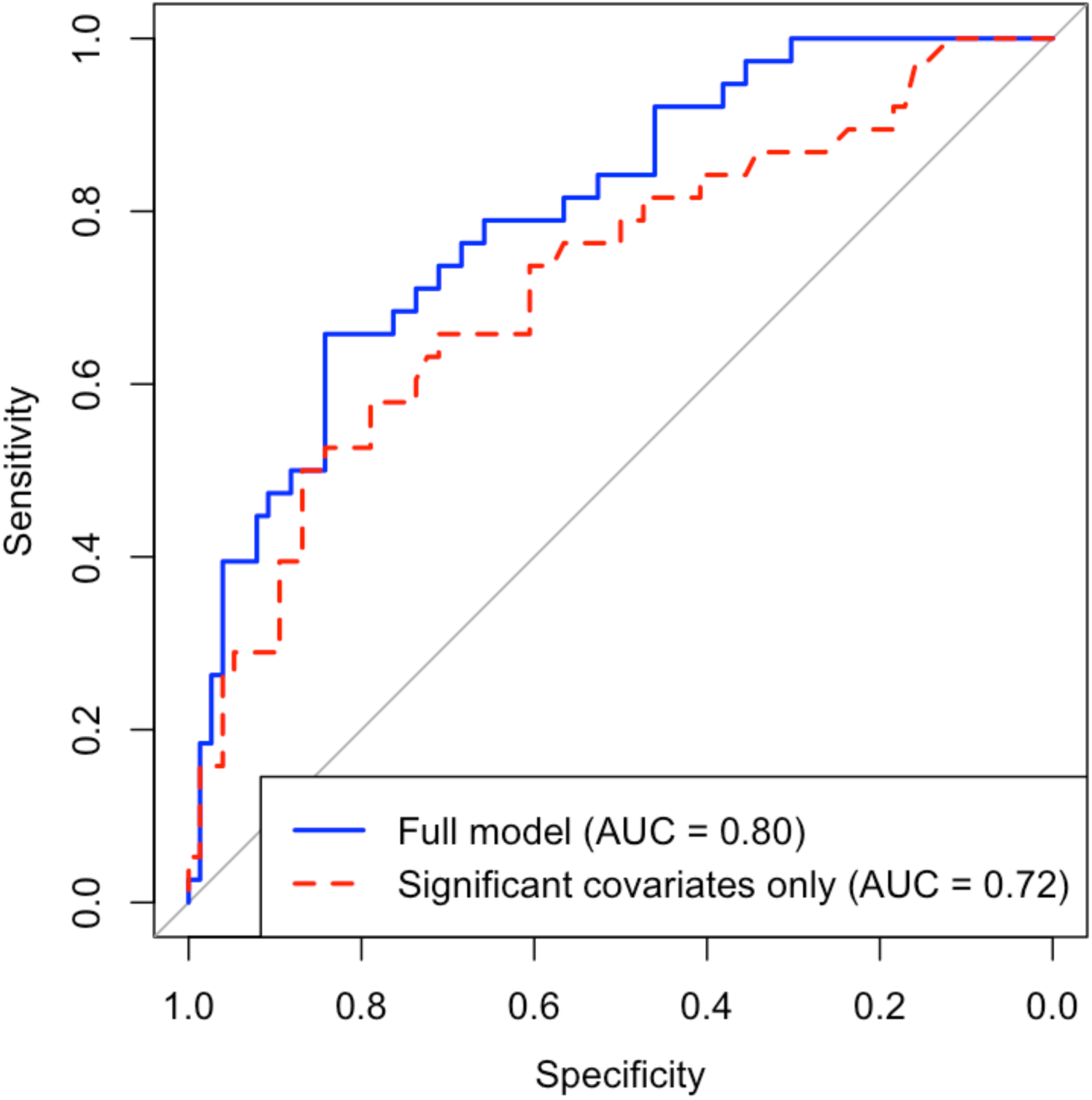
Receiver operating characteristic (ROC) curves for logistic regression models predicting *Klebsiella pneumoniae* infection. The solid blue line represents the full model including all covariates (AUC = 0.80), and the dashed red line represents the model with non-significant covariates set to their mean values (AUC = 0.72).

